# Interactions Between Cannabis Use and Chronic Pain on Sleep Architecture: Findings from In-Home EEG Recordings

**DOI:** 10.1101/2025.08.28.25334662

**Authors:** Tracy Brown, Francesca Filbey

## Abstract

**Background:** Pain and sleep disturbances are primary reasons for medicinal cannabis use. Cannabis influences both pain and sleep through its modulation of the endocannabinoid system, which regulate pain pathways and sleep regulation. Despite their interconnected roles, the effects of cannabis and chronic pain on sleep architecture are studied mainly in isolation. An integrated understanding is needed to guide use and minimize risks in this population.

**Objective:** Our primary aim was to examine the potential interactive effect of regular cannabis use on chronic pain and sleep.

**Methods:** A total of 339 nights (2,273.43 hours) of in-home sleep electroencephalogram (EEG) recordings were collected from 60 adults (50% male; 32% chronic pain; 47% cannabis use; *M*_age_= 25.25; *SE*= 1.05) over seven consecutive nights per participant. A mixed-model repeated-measures ANCOVA tested the main effects and interactions of chronic pain and regular cannabis use on total sleep time (TST), total slow-wave sleep (SWS%), total rapid-eye-movement (REM%), sleep onset latency (SOL), and number of awakenings.

**Results:** There was a significant main effect of cannabis use on SWS, TST, SOL, and REM. There was a significant main effect of chronic pain on TST. Significant interactions emerged between cannabis use and chronic pain on SWS and REM.

**Conclusions:** These findings may reflect a dysregulated sleep response in individuals using cannabis to manage chronic pain, highlighting the need to consider both beneficial and detrimental effects of cannabis on specific sleep stages.

## Introduction

The most reported reasons for medicinal cannabis use are for pain relief and improvements in sleep (1–3), with some studies reporting more than half (i.e., 64%) do so for pain (4), while others report up to 80% use it for sleep (5,6). The endocannabinoid system (ECS) plays a regulatory role in both pain and sleep (7–9). Tetrahydrocannabinol (THC), the primary psychoactive ingredient in cannabis binds with CB1 receptors in pain processing areas that lead to the reduction of nociceptive signaling (10,11), anti-inflammatory effects, and modulation of emotional response to pain (12). Neuroimaging studies indicate that THC improves top-down processing of pain by reducing activation of brain regions that contribute to the emotional response to pain (i.e., amygdala) and attention to pain (i.e., sensorimotor cortex and the dorsolateral prefrontal cortex) (13). Additionally, the pleasurable and rewarding experience associated with cannabis-induced euphoria also alleviates the negative experience of pain (14).

In terms of sleep, THC activates CB1 receptors in brain regions like the hypothalamus, amygdala, and brainstem, which are involved in sleep regulation. Cannabis use has been associated with improvements in various sleep metrics including total sleep time (TST), sleep onset latency (SOL), slow wave sleep (SWS), and number of sleep disruptions (5,15). Notably, cannabis appears to have biphasic effects on sleep where acute cannabis use shows improvements in sleep, but prolonged use reduces these benefits and potentially worsens sleep. For example, Kaul et al (2021) reported that near-daily use for three months was associated with reduced TST and SWS and increased SOL. These findings suggest non-linear, complex effects of cannabis on sleep.

Despite the role of ECS in both sleep and pain regulation (7–9), few studies have examined the combined effect of cannabis use and pain on sleep. However, in general, existing studies demonstrate sleep improvements following cannabis therapy in those with chronic pain. A THC nasal spray administered to patients with neuropathic pain resulted in improvements in subjective sleep measures (16), which is concordant to effects of synthetic THC in patients with fibromyalgia when compared to an antidepressant. In another clinical trial, smoked cannabis not only reduced pain intensity and improved subjective sleep quality in patients with neuropathic pain (17). Findings from observational studies are consistent with those from clinical trials. For example, Sznitman et al. (2020) showed that adults with chronic pain who used cannabis reported fewer night-time awakenings than those with chronic pain who did not use cannabis. Interestingly, those with more frequent use had greater wakefulness and sleep onset latency, which the authors attributed to potential cannabis tolerance (18). Taken together, existing studies show that cannabis is associated with improvements in subjective measures of sleep. However, how these findings relate to objective measures of sleep, i.e., sleep architecture, has yet to be examined.

Our review of the literature suggests that cannabis may provide a dual reinforcing effect in those with chronic pain by alleviating pain and improving sleep. However, given that improvements in sleep may diminish with prolonged use, likely through drug tolerance effects (19,20), there is a risk of escalating patterns of cannabis use. This cycle of diminished efficacy and compensatory use may heighten the risk for problematic cannabis use or the development of cannabis use disorder (CUD). The goal of this study was to address the need to understand the potential interactive relationship between regular cannabis use, pain and sleep. Given previous findings demonstrating that chronic pain and cannabis use have independent negative effects on sleep metrics, such as SWS and REM (21–24), we hypothesized an interaction between long-term cannabis use and chronic pain such that individuals with chronic pain who use cannabis will have decreased SWS and REM sleep compared to those without cannabis use and with or without chronic pain. We predicted that prolonged cannabis use would be associated with greater reductions to SWS and REM. Specifically, we will test whether long-term cannabis use leads to disrupted sleep, which in turn, contributes to reductions in sleep stages important for regulating pain.

## Methods

The Institutional Review Board of the University of Texas at Dallas approved this study.

### Participants

Sixty-two participants were recruited from the general community within the Dallas metro area via flyers and social media (i.e., Facebook, Twitter, and Reddit). The inclusion criteria were: (a) aged 18-45 years, (b) proficiency in English, and (c) ability to provide informed consent. The exclusion criteria were: (a) history of brain injury or neurological diagnosis (e.g., stroke, epilepsy, MS), (b) history of psychosis, (c) currently taking psychotropic medications, (d) pregnancy, (e) history of sleep disorder, (f) shift-work (work hours between 10 PM - 6 AM), and (g) use of illicit substance other than cannabis.

### Study Procedures and Measures

The study consisted of an initial lab visit and seven days of in-home data collection. The initial lab visit included collection of informed consent and completion of self-report questionnaires. Age and gender were collected via a demographics questionnaire. The Substance Use History Questionnaire (SUHQ) was used to assess frequency and duration of cannabis use and other substances. Chronic pain was defined based on ICD-11 (2021) definition of persistent pain for at least three months (25) as assessed using the following questions “Have you experienced persistent pain for the past 3 months or longer (i.e., chronic pain)?” and “What is the name of your pain condition. If unknown, please describe the pain you experience”.

EEG recordings were collected using the DREEM 3 headband (26). The DREEM headband contains dry electrodes positioned at frontal (F7, F8, Fp2) and occipital (O1, O2) sites in addition to sensors to monitor heart rate, respiration, and movement. DREEM’s proprietary algorithms segmented sleep into the standard stages: wake, N1, N2, N3, and REM in addition to total sleep time, sleep efficiency, sleep onset latency, and the duration of each sleep stage. DREEM’s accuracy in sleep staging has been validated against polysomnography (26).

### Data Processing and Statistical Analyses

We recorded 434 nights of sleep EEG. Of these sleep recordings, 95 nights of data were excluded due to poor data quality defined as quality score of <70% based on DREEM’s algorithm. As a result, two participants were excluded entirely from the analyses resulting in a final sample of 60 participants with 339 nights (2,273.43 hours) of usable EEG sleep recordings. Among these participants, 16 reported nociceptive pain (10 of whom with cannabis use), and 3 with cannabis use reported non-nociceptive pain (see Table 1 for sample characteristics).

**Table 1.** Demographics of study participants and ANCOVA results for differences. Gender was coded as 1 for male and 0 for female. Chronic pain was coded as “1” with pain and “0” without pain. “F”=female, “M”=male, CUDIT-R= Cannabis Use Disorder Identification Task-Revised.

|  |  |
| --- | --- |
| Total (N) | 60 |
| Gender (M/F) | 30/30 |
| Age (M/SE) | 25.25/1.05 |
| Chronic pain | 32% |
| Regular cannabis use | 47% |
| Cannabis use years (M/SE) | 3.1/0.88 |
| Cannabis use days per week (M/SE) | 3.41/0.66 |

Repeated measures analyses were performed using Linear Mixed Models (LMMs) in SPSS to examine nightly sleep metrics in relation to cannabis use and pain. The models included cannabis use (e.g., yes/no) and presence of pain (e.g., yes/no) as fixed effects, and subject ID as a random intercept to account for repeated measures nested within individuals. The dependent variables were TST, SOL, SWS, REM, and number of awakenings. To control for potential confounds, we included gender, age (ANCOVA structure). Models were estimated using restricted maximum likelihood (RML), and compound symmetry was selected as the covariance parameter.

Considering longer durations of cannabis use diminish changes to sleep architecture (5,15) and cause worse sleep in those with chronic pain (18), Pearson correlations were computed to test for the effects of years of using cannabis on sleep metrics. Data reduction was performed by averaging the sleep metrics for those reporting regular cannabis use (see Table 2).

**Table 2.** Pearson’s one-tailed correlations between years of cannabis use, pain, and sleep metrics. SOL= sleep onset latency, TST= total sleep time, SWS= slow-wave sleep, REM= rapid eye movement (\**p*<.05).

|  | *Pain* | *SOL* | *TST* | *SWS* | *REM* | *Awakenings* |
| --- | --- | --- | --- | --- | --- | --- |
| Years of<br>regular use | -.04 | .09 | .27 | **-.33\*** | -.07 | .08 |
| Pain |  | .05 | -.1 | .07 | .01 | -.1 |

## Results

The following results are based on the fixed factors and interactions between cannabis and chronic pain tested in the models. All main effects and interactions on TST, SOL, SWS, REM, and awakenings are reported in Tables 3-7.

**Table 3.** Type III fixed effects and interactions for chronic pain and cannabis use on TST.

| | *dfn* | *dfd* | *F* | *p* | $\beta$ | *SE* | 95% CI | |
| --- | --- | --- | --- | --- | --- | --- | --- | --- |
|  |  |  |  |  |  |  | Lower bound | Upper bound |
| (intercept) | 1 | 36.54 | 480.34 | <.001 | 338.85 | 25.26 | 288.49 | 389.21 |
| **Main effects** |  |  |  |  |  |  |  |  |
| Chronic pain | 1 | 44.63 | 16.84 | <.001* | 27.84 | 13.19 | **1.32** | **54.36** |
| Cannabis use | 1 | 45.95 | 6.52 | .01* | -39.21 | 16.75 | **-73.01** | **-5.42** |
| **Interaction** |  |  |  |  |  |  |  |  |
| Chronic pain<br>x<br>Cannabis use | 1 | 44.45 | 1.70 | .20 | -26.20 | 20.12 | -66.73 | 14.33 |
| **Covariates** |  |  |  |  |  |  |  |  |
| Sex | 1 | 46.40 | 2.00 | .16 | 12.92 | 9.13 | -5.47 | 31.30 |
| Age | 1 | 51.19 | 2.68 | .11 | -16.70 | 10.20 | -37.17 | 3.77 |

**Table 4.**
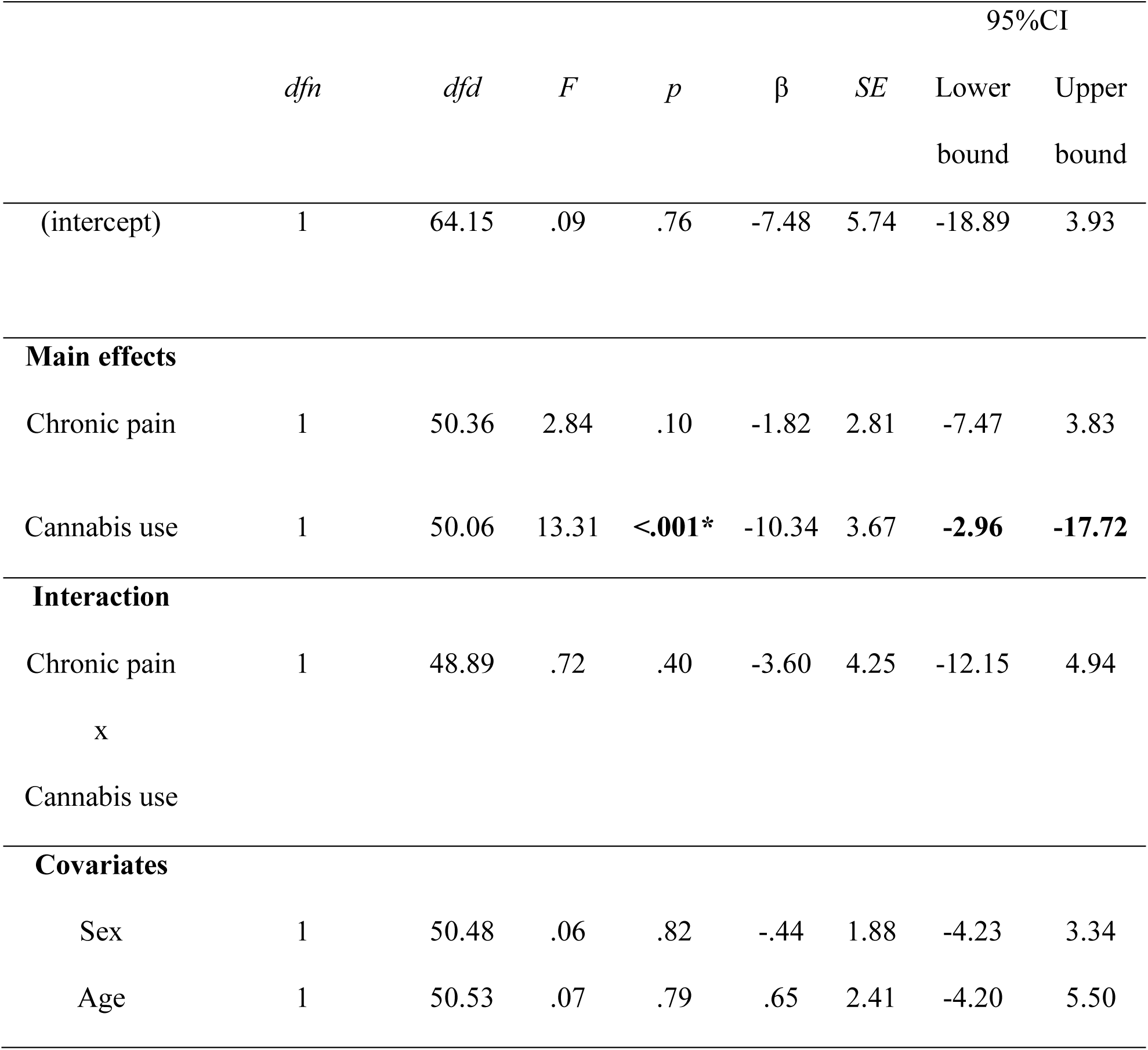
Type III fixed effects and interactions for chronic pain and cannabis use on SOL.

**Table 5.**
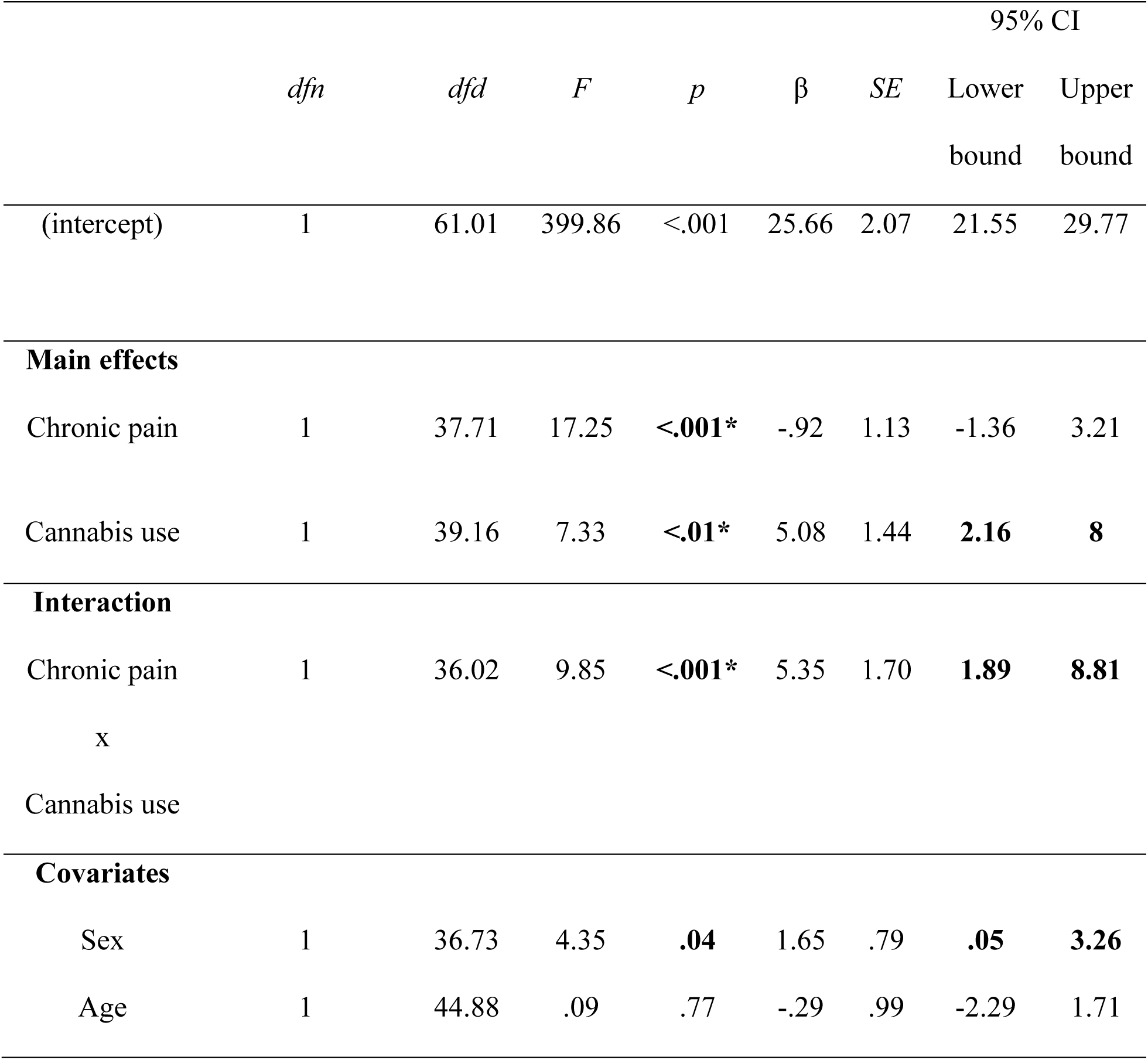
Type III fixed effects and interactions for chronic pain and cannabis use on SWS.

|  |  |  |  |  |  |  | 95% CI |  |
| --- | --- | --- | --- | --- | --- | --- | --- | --- |
| | *dfn* | *dfd* | *F* | *p* | $\beta$ | *SE* | Lower bound | Upper bound |
| (intercept) | 1 | 61.01 | 399.86 | <.001 | 25.66 | 2.07 | 21.55 | 29.77 |
| **Main effects** |  |  |  |  |  |  |  |  |
| Chronic pain | 1 | 37.71 | 17.25 | <.001* | -.92 | 1.13 | -1.36 | 3.21 |
| Cannabis use | 1 | 39.16 | 7.33 | <.01* | 5.08 | 1.44 | **2.16** | **8** |
| **Interaction** |  |  |  |  |  |  |  |  |
| Chronic pain<br>x<br>Cannabis use | 1 | 36.02 | 9.85 | <.001* | 5.35 | 1.70 | **1.89** | **8.81** |
| **Covariates** |  |  |  |  |  |  |  |  |
| Sex | 1 | 36.73 | 4.35 | **.04** | 1.65 | .79 | **.05** | **3.26** |
| Age | 1 | 44.88 | .09 | .77 | -.29 | .99 | -2.29 | 1.71 |

**Table 6.** Type III fixed effects and interactions for chronic pain and cannabis use on REM.

|  |  |  |  |  |  |  | 95% CI |  |
| --- | --- | --- | --- | --- | --- | --- | --- | --- |
| | *dfn* | *dfd* | *F* | *p* | $\beta$ | *SE* | Lower bound | Upper bound |
| (intercept) | 1 | 89.89 | 256.03 | <.001 | 19.57 | 2.40 | 14.82 | 24.33 |
| **Main effects** |  |  |  |  |  |  |  |  |
| Chronic pain | 1 | 50.41 | 11.14 | <.001* | 1.87 | 1.36 | -.87 | 4.60 |
| Cannabis use | 1 | 53.96 | 8.01 | <.01* | -8.50 | 1.83 | **-12.18** | **-4.83** |
| **Interaction** |  |  |  |  |  |  |  |  |
| Chronic pain<br>x<br>Cannabis use | 1 | 49.04 | 25.19 | <.001* | -10.73 | 2.14 | **-15.03** | **-6.43** |
| **Covariates** |  |  |  |  |  |  |  |  |
| Sex | 1 | 52.33 | 2.12 | .15 | -1.38 | .95 | -3.28 | .52 |
| Age | 1 | 53.06 | .17 | .68 | .46 | 1.13 | -1.80 | 2.73 |

**Table 7.** Type III fixed effects and interactions for chronic pain and cannabis use on awakenings.

|  |  |  |  |  |  |  | 95% CI |  |
| --- | --- | --- | --- | --- | --- | --- | --- | --- |
| | *dfn* | *dfd* | *F* | *p* | $\beta$ | *SE* | Lower bound | Upper bound |
| (intercept) | 1 | 68.27 | 140.09 | <.001 | 20.08 | 2.38 | 15.36 | 24.80 |
| **Main effects** |  |  |  |  |  |  |  |  |
| Chronic pain | 1 | 45.60 | .60 | .44 | 2.31 | 1.33 | -4.98 | .36 |
| Cannabis use | 1 | 47.80 | 1.68 | .20 | .08 | 1.82 | -3.75 | 3.59 |
| **Interaction** |  |  |  |  |  |  |  |  |
| Chronic pain<br>x<br>Cannabis use | 1 | 45.74 | 2.00 | .16 | 3.00 | 2.12 | -1.27 | 7.27 |
| **Covariates** |  |  |  |  |  |  |  |  |
| Sex | 1 | 46.56 | .18 | .67 | .38 | .90 | -1.43 | 2.20 |
| Age | 1 | 52.09 | .02 | .89 | -.16 | 1.16 | -2.49 | 2.16 |

### Main Effects of Cannabis on Sleep Metrics

The significant main effects of regular cannabis included: (1) lower TST (β= -39.21, 95%; CI [-73.01, -5.42]), (2) decreased SOL (β= -10.34, 95%; CI [-2.96, -17.72]), (3) increased SWS (β= 5.08, 95%CI [2.16, 8]), and (4) less REM sleep (β= -8.5, 95%CI [-12.18,-4.83]). There was no significant effect of cannabis on awakenings (β= 0.08, 95%CI [-3.75, 3.59]).

### Main Effects of Chronic pain on Sleep Metrics

There was a significant main effect of pain on TST such that chronic pain was associated with greater TST (β= 27.84, 95%CI [1.32, 54.36]). No significant main effect of chronic pain was found on SOL (β= -1.82, 95%CI [-7.47, 3.83]), SWS (β= -0.92, 95%CI [-1.36, 3.21]), REM (β= 1.87, 95%CI [-0.87, 4.6]), or awakenings (β= 2.31, 95%CI [-4.98, 0.36]).

### Interactions of Cannabis and Chronic pain on Sleep Metrics

There was a significant interaction between cannabis use and pain on: (1) SWS (β= 5.35, 95%CI [1.89, 8.81]) such that cannabis use and chronic pain led to greater SWS (see Figure 1); and (2) REM sleep (β= -10.73, 95%CI [-15.03, -6.43]) such that chronic pain and cannabis use both decreased amounts of REM sleep (see Figure 2). No significant interactions were found between cannabis use and pain on TST (β= -26.2, 95%CI [-66.73, 14.33]), SOL (β= -3.6, 95%CI [-12.15, 4.94]), or awakenings (β= 2.12, 95%CI [-1.27, 7.27]).

**Figure 1.**
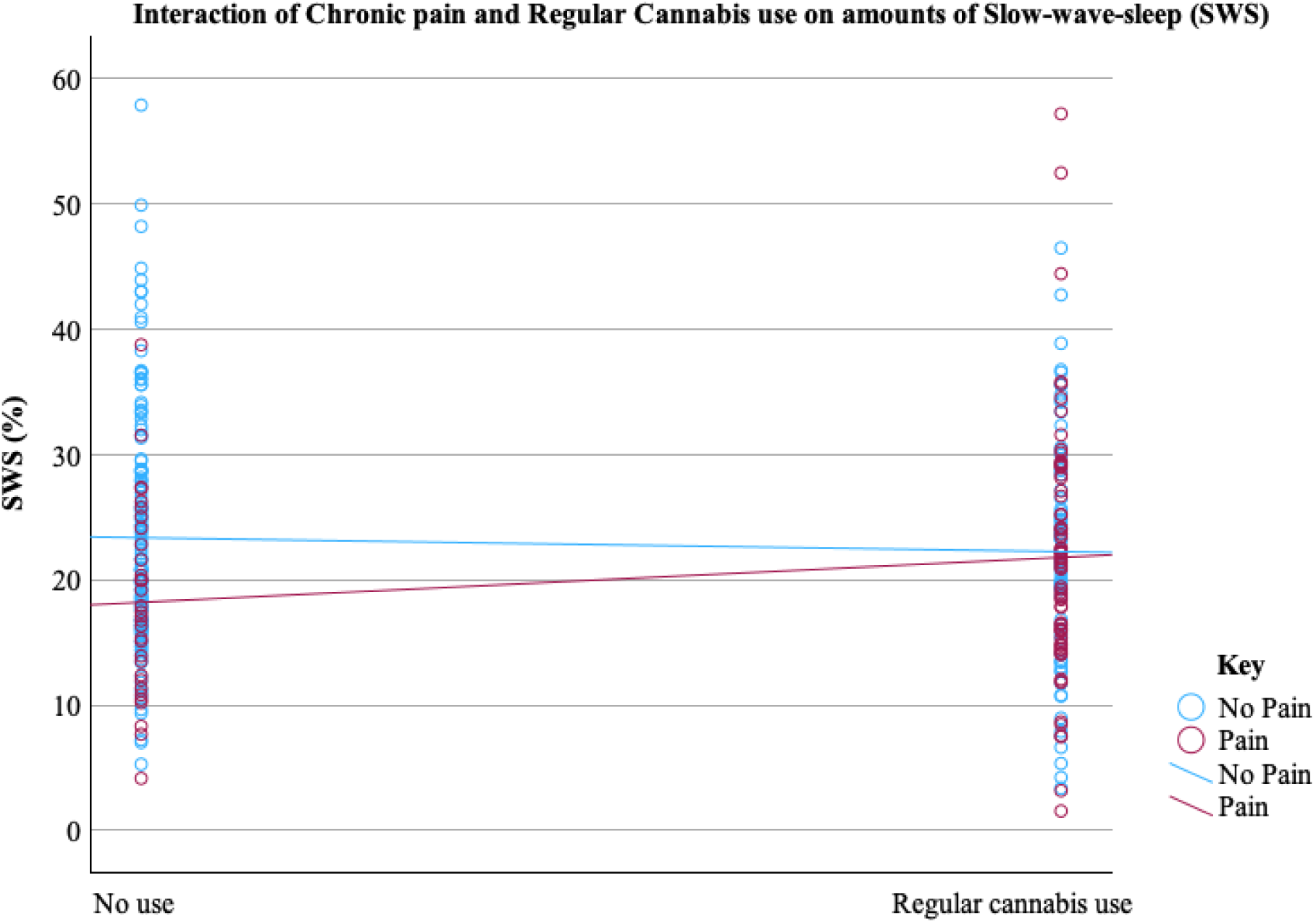
The interaction of regular cannabis use and chronic pain on slow-wave sleep (SWS).

**Figure 2.**
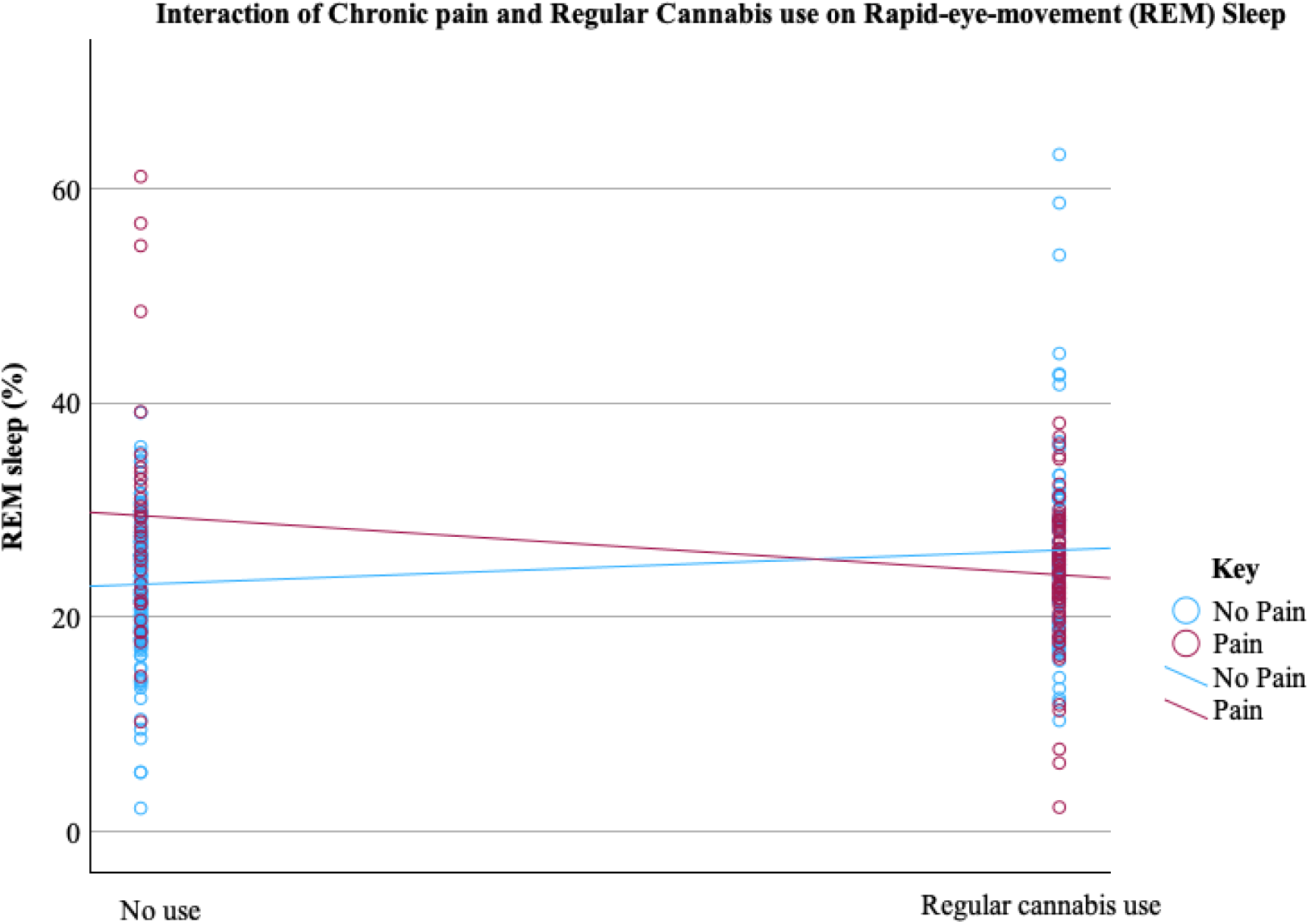
The interaction of regular cannabis use and chronic pain on rapid-eye-movement (REM) sleep.

### Correlations between Duration of Cannabis use and Sleep Metrics

Results from the one-tailed Pearson correlations revealed a significant negative association between years of regular cannabis use and slow-wave sleep (SWS), such that more years of use was associated with reduced SWS (p=.04). There were no significant correlations between years of regular cannabis use and TST (p=.12), SOL (p=.32), awakenings (p=.38), or REM sleep (p=.36).

### Post-hoc Analyses

We conducted linear mixed regression analyses to examine potential effects of type of pain on our outcomes. Our findings showed no significant effect of pain type (i.e., nociceptive vs. non-nociceptive pain types) on TST (β= -1.71, 95%CI [-87.71, 84.29]), SOL (β= -8.84, 95%CI [-25.19, 7.51]), SWS (β= -6.37, 95%CI [-13.31, 0.56]), REM sleep (β= -2.78, 95%CI [-12.43, 6.88]), or awakenings (β= 4.93, 95%CI [-2.88, 12.73]).

## Discussion

The interactive effect of cannabis use and chronic pain on sleep remains unclear despite the acknowledged bi-directional relationship between pain and sleep (i.e., poor sleep increases pain sensitivity, and more pain worsens sleep) and the regulatory role of the endocannabinoid system on both. Our findings indicated that the combination of regular cannabis use and chronic pain was associated with increased SWS and decreased REM, suggesting that the effects of cannabis use on SWS and REM differ depending on the presence of chronic pain.

The interactive effect of cannabis and pain on sleep reflects the complex overlapping roles of the ECS in regulating pain, sleep, and their shared neurobiological pathways (27). Specifically, THC interacts with CB1 receptors that are densely located in brain areas involved in both nociception and sleep regulation, such as the hypothalamus, amygdala, thalamus, and brainstem (8). Studies suggest that THC attenuates pain signaling and inhibits GABA through the activation of CB1 receptors and indirectly by increasing anandamide (AEA) and 2-arachidonoylglycerol (2-AG) that complete retrograde synaptic signaling of CB1 receptors (28,29). This increase of AEA and 2-AG from THC also promotes sleep (28–31). However, in individuals with chronic pain, the ECS is dysregulated, which could cause exogenous cannabinoids like THC to alter sleep architecture differently than in those without pain. Given that immune cell activation is reduced during SWS, (Ranjbaran et al., 2007), increasing the amount of SWS may therefore provide better pain outcomes (33). This suggests a negative feedback loop between sleep architecture and pain, such that pain increases sleep disruptions, decreases SWS, and triggers immune cell activation, which leads to increased inflammation and experienced pain (34,35). Thus, the direct pain-alleviating qualities of THC (5,29,36) and indirect pain alleviation via increased SWS may benefit in regulating pain.

The complex interaction between cannabis use and chronic pain in shaping SWS and REM suggests a potential tradeoff whereby increased SWS might be at the cost of impairing REM. SWS is critical for physical restoration and immune function (32), so this increase may reflect a compensatory mechanism to recover from the physiological strain of chronic pain, enhanced by cannabis. On the other hand, a marked reduction in REM sleep could indicate a disruption in cognitive or emotional restorative processes, which may have long-term implications for mental health and pain perception (37,38). For example, reduced REM sleep impairs the integration of newly acquired non-declarative, procedural, and emotional memories (39) and compromises emotional regulatory mechanisms that contribute to heightened emotional reactivity and difficulty in coping with stressors (40). Furthermore, fMRI findings show that reduced REM sleep increases amygdala activation and medial prefrontal cortex control during emotional tasks (37), which is similar to the functional connectivity dysregulation reported in individuals with chronic pain at rest (41).

Although our results of increased SWS and reduced REM sleep in the current sample of long-term cannabis users with chronic pain is consistent with findings of acute effects of cannabis on sleep (42), our correlational analyses revealed that greater years of regular cannabis use was associated with decreased SWS across all participants. This latter finding is consistent with evidence of tolerance and receptor downregulation observed in long-term users (21,43), suggesting that while cannabis may initially enhance SWS, these benefits diminish with chronic exposure. This finding provides further evidence for the biphasic effect of cannabis on sleep, where acute cannabis use promotes sleep through direct ECS modulation, while chronic use leads to tolerance, receptor downregulation, and disrupted sleep architecture. The paradoxical finding of reduced SWS with greater duration of cannabis use across all participants (i.e., those with pain and without pain) with observations of increased SWS in those with chronic pain may indicate that cannabis continues to provide some benefit for sleep in chronic pain patients through pain relief, even as tolerance develops to its direct effects on the ECS. By contrast, in individuals without chronic pain, cannabis effects on sleep are more direct, allowing the biphasic pattern of initial benefit (increased SWS with short-term use) followed by tolerance-related disruption (reduced SWS with long-term use) to emerge more clearly. This suggests that pain-related sleep disturbances may mask cannabis-related changes in sleep architecture. Our findings indicating no effect of pain type on our outcomes support previous reports of similar sleep disturbances across different pain diagnoses (44).

### Limitations and Conclusions

Interpretation of these findings is limited by the study’s cross-sectional design. A longitudinal approach is needed to understand the potential parabolic function of cannabis use in populations with chronic pain. This knowledge could inform clinical monitoring of patients using cannabis therapeutically.

To conclude, these findings suggest a complex interaction between cannabis use and chronic pain in shaping sleep architecture, particularly in SWS and REM sleep. Our findings suggest that the interaction between cannabis and pain leads to a potential “trade-off” such that the beneficial sleep process is enhanced (SWS) while another critical function served by REM sleep may be compromised. Cannabis use in people with chronic pain may promote more deep, physically restorative sleep (SWS), but at the functional cost of reduced REM sleep, which could impair emotional and cognitive health over time. This trade-off could paradoxically worsen the long-term experience of pain, mood, or stress, despite short-term sleep improvements.

## Data Availability

All data produced in the present study are available upon reasonable request to the authors

